# Comparative Analysis of CHEK1, Extrachromosomal DNA Dynamics, and 1p/19q Status in Lower-Grade and High-Grade Glioma

**DOI:** 10.1101/2025.10.02.25337199

**Authors:** Steven Lehrer, Peter Rheinstein

## Abstract

**Background:** Extrachromosomal DNA (ecDNA) drives rapid tumor evolution, genomic instability, and therapy resistance in gliomas by amplifying oncogenes and generating intratumoral heterogeneity. These genomic alterations induce replication–transcription conflicts, creating a dependence on checkpoint kinase 1 (CHK1), encoded by *CHEK1*, to resolve replication stress.

**Methods:** We analyzed genomic, transcriptomic, and clinical data from The Cancer Genome Atlas (TCGA) lower-grade glioma (LGG) and glioblastoma (GBM) cohorts using UCSC Xena and cBioPortal. *CHEK1* expression, copy number variation, mutation burden, and fraction of genome altered were correlated with overall survival, *TP53*, and *ATRX* status, and *1p/19q* co-deletion. Kaplan–Meier survival analyses, log-rank tests, and co-occurrence analyses were performed, and correlations with genomic instability were assessed.

**Results:** High *CHEK1* expression and amplification were significantly associated with reduced survival, particularly in *1p/19q*-non-codeleted LGG (P < 0.001). *CHEK1* amplification correlated with increased mutation burden and a higher fraction of genome altered, indicating a link between replication stress signaling and chromosomal instability. Co-occurrence analysis revealed strong associations between *CHEK1* alterations and *TP53* mutations, highlighting their cooperative role in replication stress responses. In contrast, *1p/19q*-codeleted tumors exhibited lower *CHEK1* expression and improved prognosis, suggesting reduced replication stress in this molecular subtype.

**Conclusions:** *CHEK1* amplification and overexpression are hallmarks of ecDNA-driven gliomas, linking replication stress to poor clinical outcomes. *CHEK1* status—particularly when integrated with *1p/19q* co-deletion and genomic instability metrics—may serve as a prognostic biomarker and a predictive marker for therapeutic strategies targeting replication stress. These findings support the development of CHK1 inhibitors and combination approaches as precision treatments for genomically unstable gliomas. A subset of patients (1p/19q-non-codeleted LGG) where CHEK1 expression/amplification is present may have a prognostic and predictive marker for replication stress-targeted therapies.

## Introduction

Gliomas are the most common primary malignant tumors of the central nervous system and remain among the most therapeutically challenging cancers. Despite advances in molecular classification, including the integration of *IDH* mutation status and *1p/19q* co-deletion into diagnostic criteria, prognosis remains poor, particularly for glioblastoma (GBM), the most aggressive form of the disease. A key driver of this treatment resistance is the extraordinary genomic plasticity of gliomas, which enables rapid adaptation under selective pressure and the emergence of therapy-resistant clones [1].

One of the most important recent discoveries in cancer biology is the role of extrachromosomal DNA (ecDNA)—circular, gene-rich DNA elements that exist outside the canonical chromosomal framework—in driving this plasticity [2-4]. Unlike chromosomal amplification, which follows Mendelian inheritance patterns, ecDNA replicates and segregates unpredictably during mitosis. This stochastic inheritance creates profound intratumoral heterogeneity and accelerates tumor evolution. EcDNA frequently carries oncogenes such as *EGFR* and *MYC* and regulatory or immunomodulatory elements that enhance tumor growth and immune evasion. It also promotes catastrophic genomic rearrangements that allow cancer cell populations to rapidly reconfigure their genetic landscape in response to therapy.

A critical consequence of ecDNA-driven oncogene amplification is the induction of replication-transcription conflicts [5]. Because both transcriptional and replication machinery traverse the same DNA template, their collision generates replication stress—an intrinsic vulnerability of ecDNA-positive tumors. To resolve this stress and maintain genome integrity, tumor cells depend heavily on the checkpoint kinase 1 (CHK1) pathway, encoded by the *CHEK1* gene [6]. CHK1 is a serine/threonine kinase central to the DNA damage response (DDR), coordinating cell cycle arrest and replication fork stabilization. Its heightened activation in ecDNA-driven cancers suggests that *CHEK1* amplification or overexpression could serve not only as a biomarker of replication stress but also as a therapeutic vulnerability.

Importantly, the genomic context—particularly *1p/19q* co-deletion status—might modulate the impact of *CHEK1* on glioma biology [7]. Tumors with *1p/19q* co-deletion, characteristic of oligodendrogliomas, could show less genomic instability and improved outcomes, whereas *1p/19q*-intact astrocytomas could exhibit higher *CHEK1* expression and worse prognosis. This possible dichotomy underscores the need for stratified analyses that consider molecular subtypes when evaluating replication stress pathways as potential therapeutic targets.

The present study builds on these insights by systematically analyzing *CHEK1* expression, copy number variation, and genomic instability across lower-grade glioma (LGG) and GBM cohorts from The Cancer Genome Atlas (TCGA). We further examine how *1p/19q* co-deletion status influences the prognostic significance of *CHEK1*, and whether its activation correlates with mutational burden and chromosomal chaos— hallmarks of ecDNA-driven tumor evolution. By linking *CHEK1* status to clinical outcomes and genomic features, we aim to identify patient subgroups most likely to benefit from therapies targeting replication stress responses.

## Methods

### Data Acquisition and Cohorts

We conducted a retrospective bioinformatic analysis of genomic and clinical data from *The Cancer Genome Atlas* (TCGA) lower-grade glioma (LGG) and glioblastoma (GBM) cohorts. All data were accessed from publicly available repositories and included only de-identified patient information. Gene expression (RNA-seq, FPKM-UQ), copy number variation (CNV), somatic mutation data, and clinical annotations (including overall survival, histopathological diagnosis, and molecular subtype) were downloaded via the UCSC Xena Browser (https://xenabrowser.net) [8]. Additional analyses of mutual exclusivity, co-occurrence, and genomic alteration frequencies were performed using cBioPortal for Cancer Genomics (https://www.cbioportal.org) [9].

### Molecular and Genomic Variables

We focused on the following molecular variables:

- CHEK1 expression: Quantified as logL(FPKM-UQ + 1) and stratified into high vs. low groups using cohort-specific median or quartile cutoffs.
- Copy number status: Defined using DNAcopy segmentation and categorized as deletion, diploid, or amplification.
- Mutation burden: Total nonsynonymous mutations per tumor.
- Fraction of genome altered (FGA): Percentage of the genome affected by CNV, used as a surrogate for genomic instability.
- Molecular subtype markers: *TP53* and *ATRX* mutation status, and *1p/19q* co-deletion.

### Survival and Statistical Analyses

Overall survival (OS) was analyzed using Kaplan–Meier curves, and differences between groups were assessed using the log-rank test. Hazard ratios (HRs) with 95% confidence intervals (CIs) were estimated using Cox proportional hazards models. Correlations between *CHEK1* status and continuous variables (mutation burden, FGA) were assessed using Spearman’s rank and Pearson’s correlation coefficients as appropriate.

### Co-occurrence and Exclusivity Analysis

To identify statistically significant co-occurring genomic events, we performed pairwise analysis of *CHEK1, TP53*, and *ATRX* mutations using Fisher’s exact test. Odds ratios (ORs), p-values, and Benjamini-Hochberg–adjusted q-values were computed to control for false discovery rate (FDR) [10]. These analyses contextualized *CHEK1* alterations within established glioma molecular pathways.

### Results

CHEK1 Expression Predicts Survival Across Glioma Grades (Figure 1). In LGG, high CHEK1 expression (≥1.033) was significantly associated with poor survival (P < 0.001).

**Figure 1.**
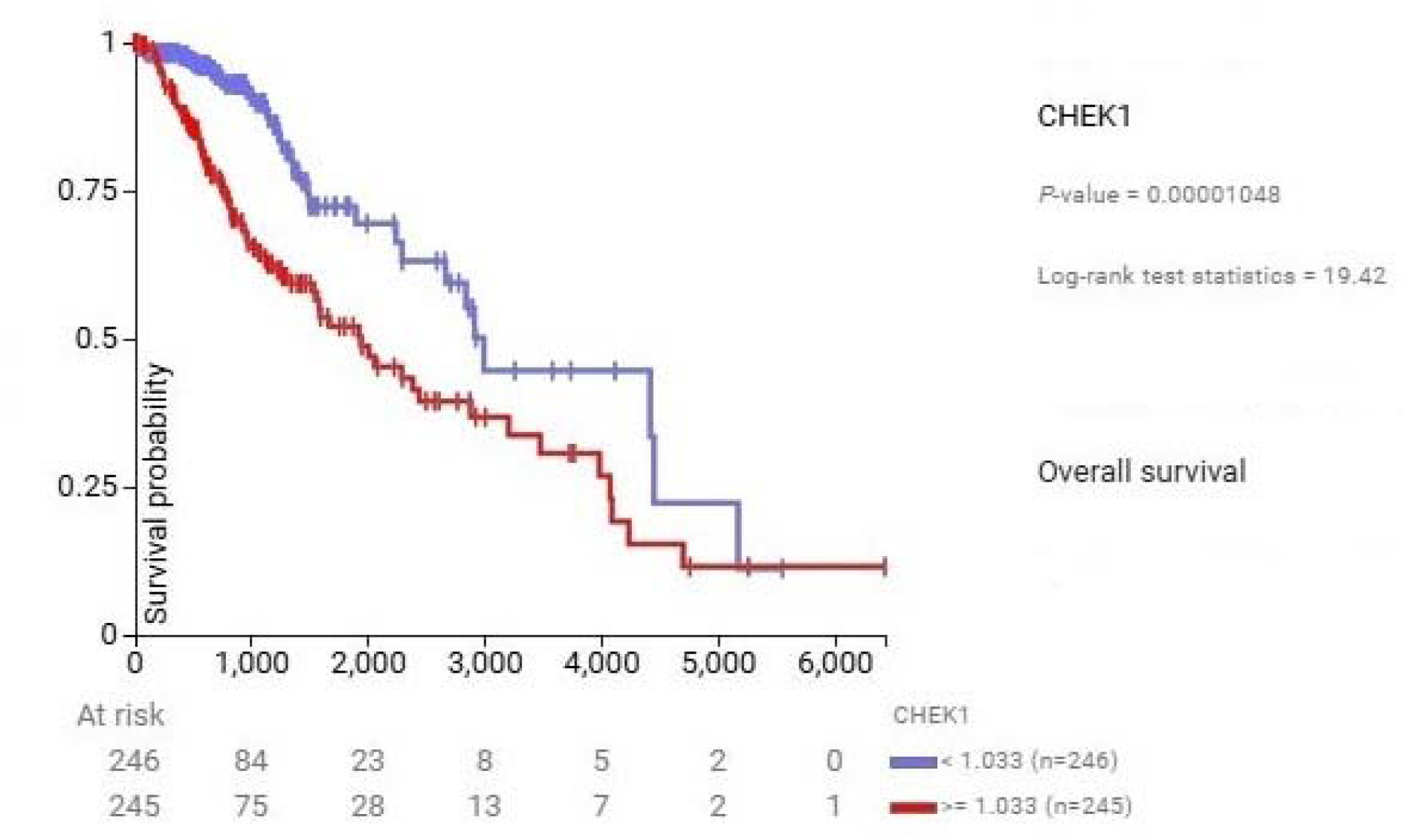
Kaplan–Meier survival curve showing overall survival in TCGA LGG stratified by CHEK1 expression.

In GBM (Figure 2), CHEK1 expression showed a similar trend but with less prognostic separation (p = 0.025), likely reflecting saturation of ecDNA-driven stress pathways. CHEK1 Dependence Is More Pronounced in LGG (Figure 1). CHEK1 dependence emerges earlier in tumorigenesis and serves as a stronger prognostic marker in LGG. In GBM, pervasive ecDNA activity leads to uniform CHEK1 activation, reducing survival discrimination.

**Figure 2.**
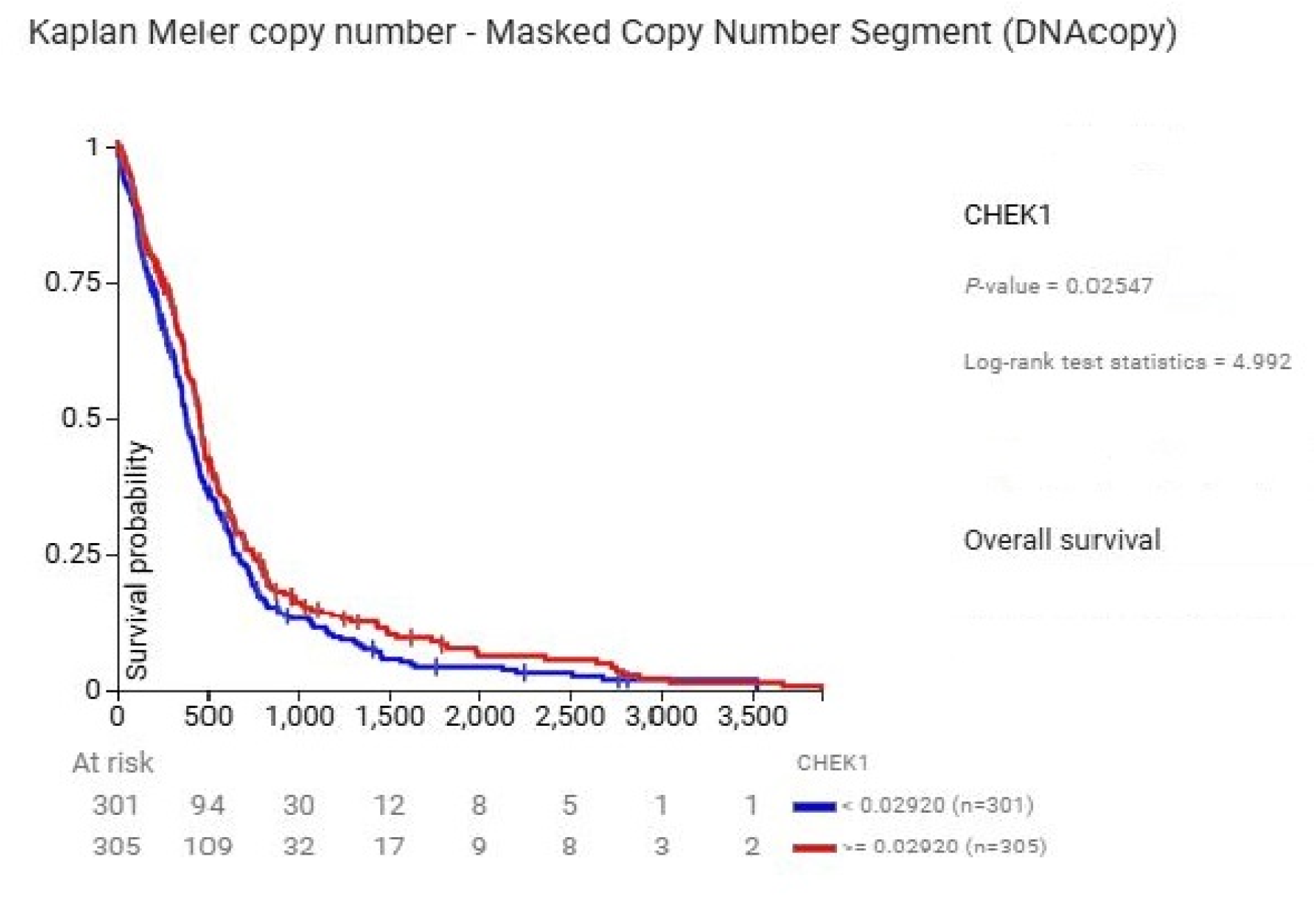
Kaplan–Meier survival curve for TCGA GBM masked copy number segments stratified by CHEK1 expression.

1p/19q Co-Deletion Modulates CHEK1 Prognostic Impact (Figure 3). 1p/19q codeleted tumors exhibited significantly lower CHEK1 expression and improved survival, even in the presence of copy number variant (CNV) gains. Non-codeleted astrocytomas displayed higher CHEK1 levels and worse outcomes, suggesting greater ecDNA-driven replication stress in these subtypes, which use the older classification [11].

**Figure 3.**
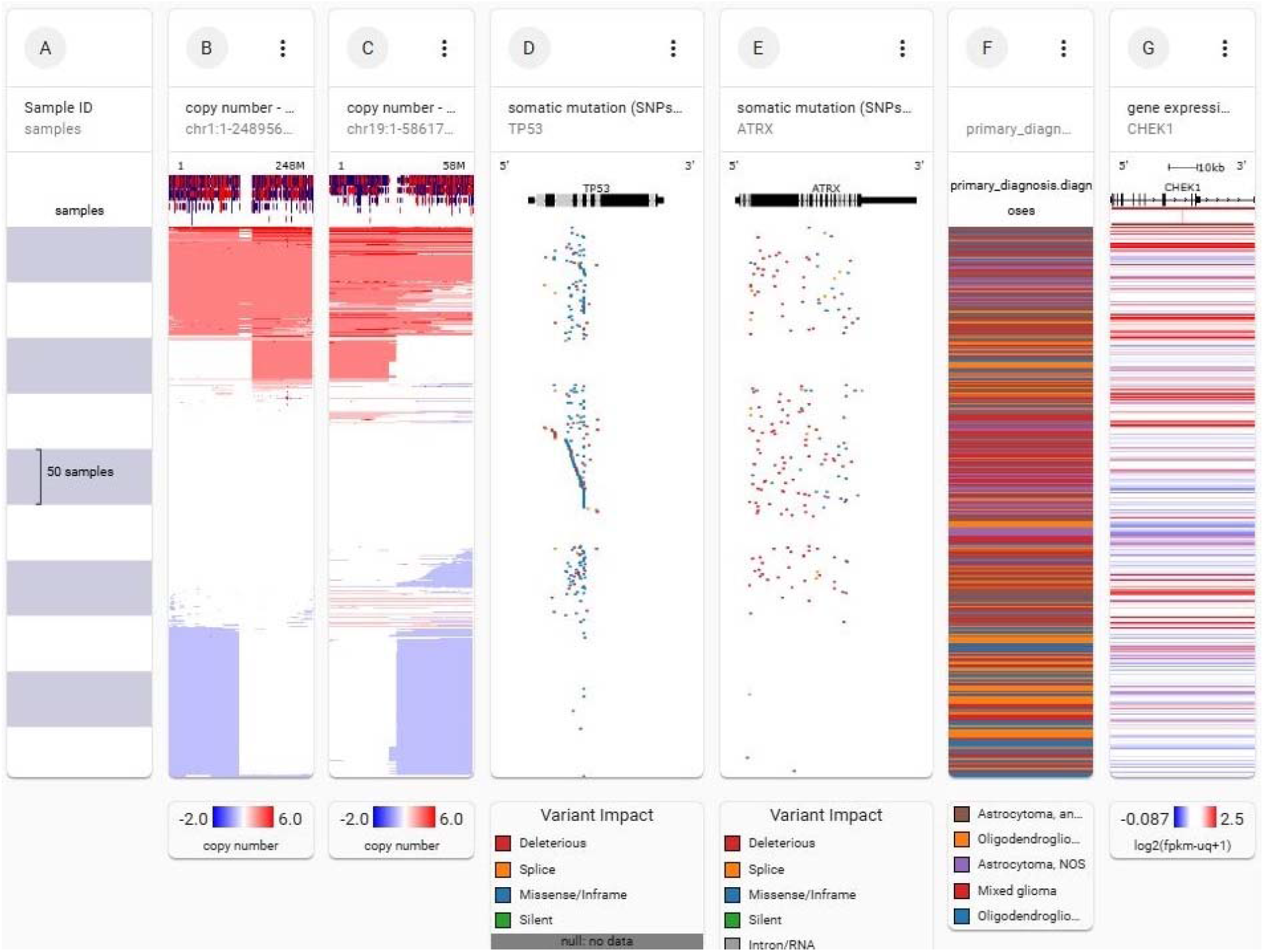
Integrated genomic landscape of CHEK1 in TCGA lower-grade glioma (LGG) with emphasis on 1p/19q status, TP53/ATRX mutations, and primary tumor subtype. This multi-panel heatmap from the UCSC Xena Browser shows the integrated copy number, mutation, and gene expression profiles of 495 LGG samples from TCGA, highlighting the relationship between CHEK1 activity, genomic alterations, and molecular subtypes. (A) Sample identifiers for all tumors in the cohort. (B–C) Copy number variation (CNV) across chromosome arms 1p and 19q. Co-deletion of these regions — visible as concurrent deep blue losses — is a defining hallmark of oligodendroglioma and is associated with favorable prognosis. (Tumors with 1p/19q co-deletion represented by vertical blue bars.) Tumors without 1p/19q co-deletion typically show partial or absent loss (red/neutral regions) and exhibit more aggressive clinical behavior. (D–E) Somatic mutation landscapes for TP53 and ATRX, two canonical astrocytoma-associated genes. TP53 mutations (predominantly deleterious and missense) and ATRX alterations (including truncating and splice variants) are enriched in 1p/19q–non-codeleted astrocytomas, indicating a mutually exclusive pattern with oligodendrogliomas. (F) Primary histopathological diagnosis for each case, illustrating the strong correspondence between molecular features and WHO classification: oligodendrogliomas cluster with 1p/19q co-deletion, whereas astrocytomas and mixed gliomas are associated with TP53/ATRX alterations and genomic instability. (G) Gene expression heatmap of CHEK1 (logL[FPKM-UQ + 1]). High CHEK1 expression (red) is significantly enriched in non-1p/19q codeleted astrocytomas and correlates with copy number gain, TP53/ATRX mutations, and increased genomic instability. In contrast, oligodendrogliomas with 1p/19q co-deletion exhibit consistently low CHEK1 expression (blue), reflecting reduced replication stress and improved clinical outcomes. Together, these data demonstrate that CHEK1 amplification and overexpression are hallmarks of 1p/19q–non-codeleted gliomas with elevated replication stress and genomic instability. This molecular context underscores CHEK1’s role as a prognostic biomarker and therapeutic vulnerability in ecDNA-driven tumor evolution

We examined the relationship between CHEK1 copy-number status and global genomic instability in TCGA lower-grade gliomas. As shown in Figure 4, tumors with CHEK1 amplification displayed a higher fraction of genome altered and elevated mutational burden compared to diploid or deleted cases. Spearman correlation analysis revealed a significant positive association between genomic instability and mutation count (ρ = 0.51, p = 3.17×10−^3^L), consistent with a replication-stress–driven feedback loop. These data suggest that CHEK1 activation is tightly coupled to the genomic chaos characteristic of ecDNA-positive tumors, underscoring its potential as both a biomarker and therapeutic target.

**Figure 4.**
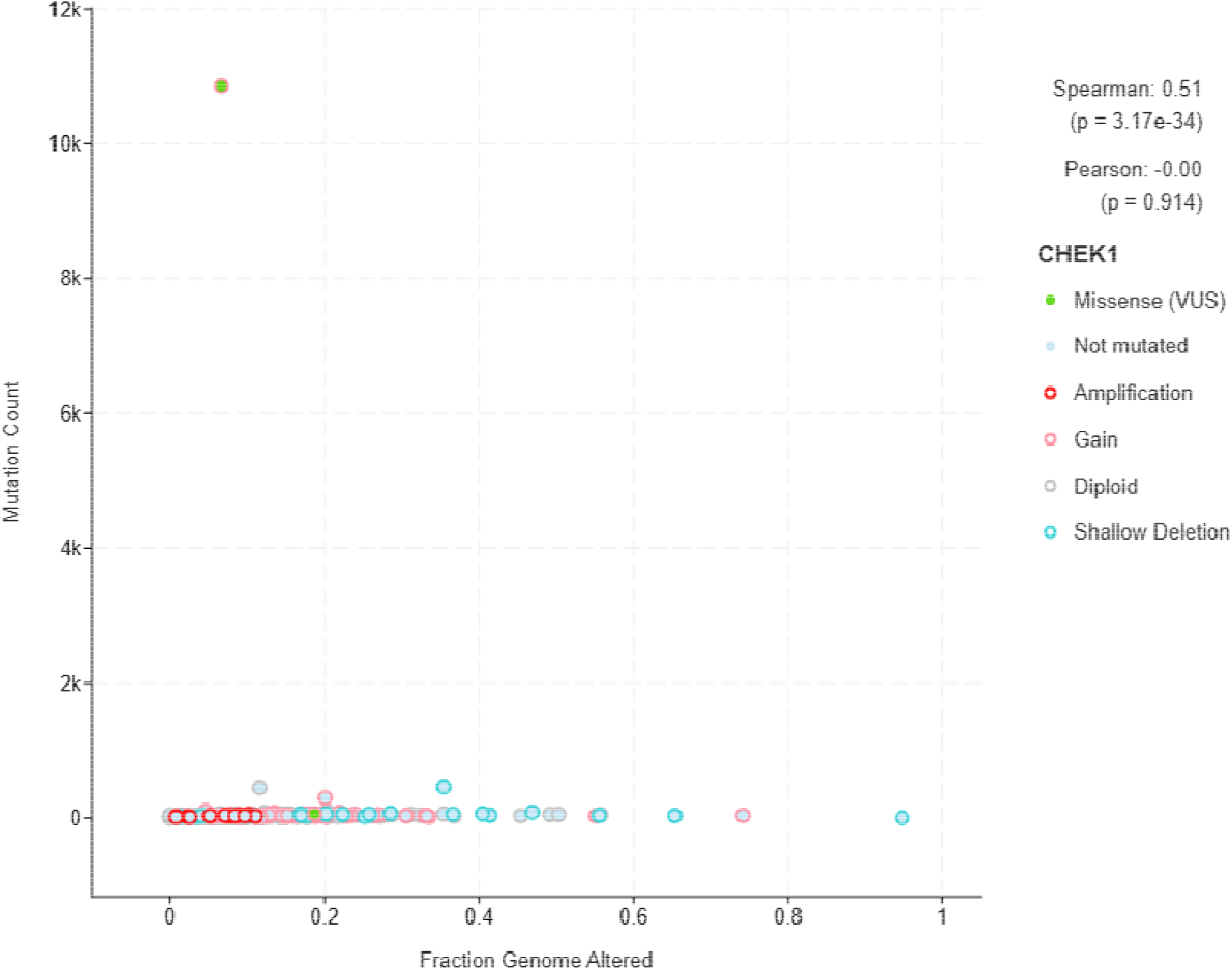
Scatter plot showing mutation count versus fraction of genome altered, stratified by CHEK1 status in TCGA lower-grade glioma. CHEK1 amplification is enriched in highly altered, mutation-rich tumors (Spearman ρ = 0.51, p = 3.17×10;^−3^).

Figure 5 is a scatter that shows the association between total mutation count and fraction of genome altered in TCGA glioblastoma (GBM) samples, stratified by *CHEK1* mutation and copy number status. Each point represents an individual tumor sample, colored by *CHEK1* alteration type (truncating mutation, missense variant of uncertain significance, gain, diploid, or shallow deletion). Unlike lower-grade gliomas, *CHEK1* status in GBM shows no significant correlation with either mutation burden or genomic instability (Spearman ρ = 0.06, *p* = 0.246; Pearson *r* ≈ 0, *p* = 0.974). These findings suggest that in advanced disease, replication stress and chromosomal chaos are pervasive and decoupled from *CHEK1* alterations, reflecting widespread genomic instability and checkpoint pathway saturation characteristic of late-stage gliomas.

**Figure 5.**
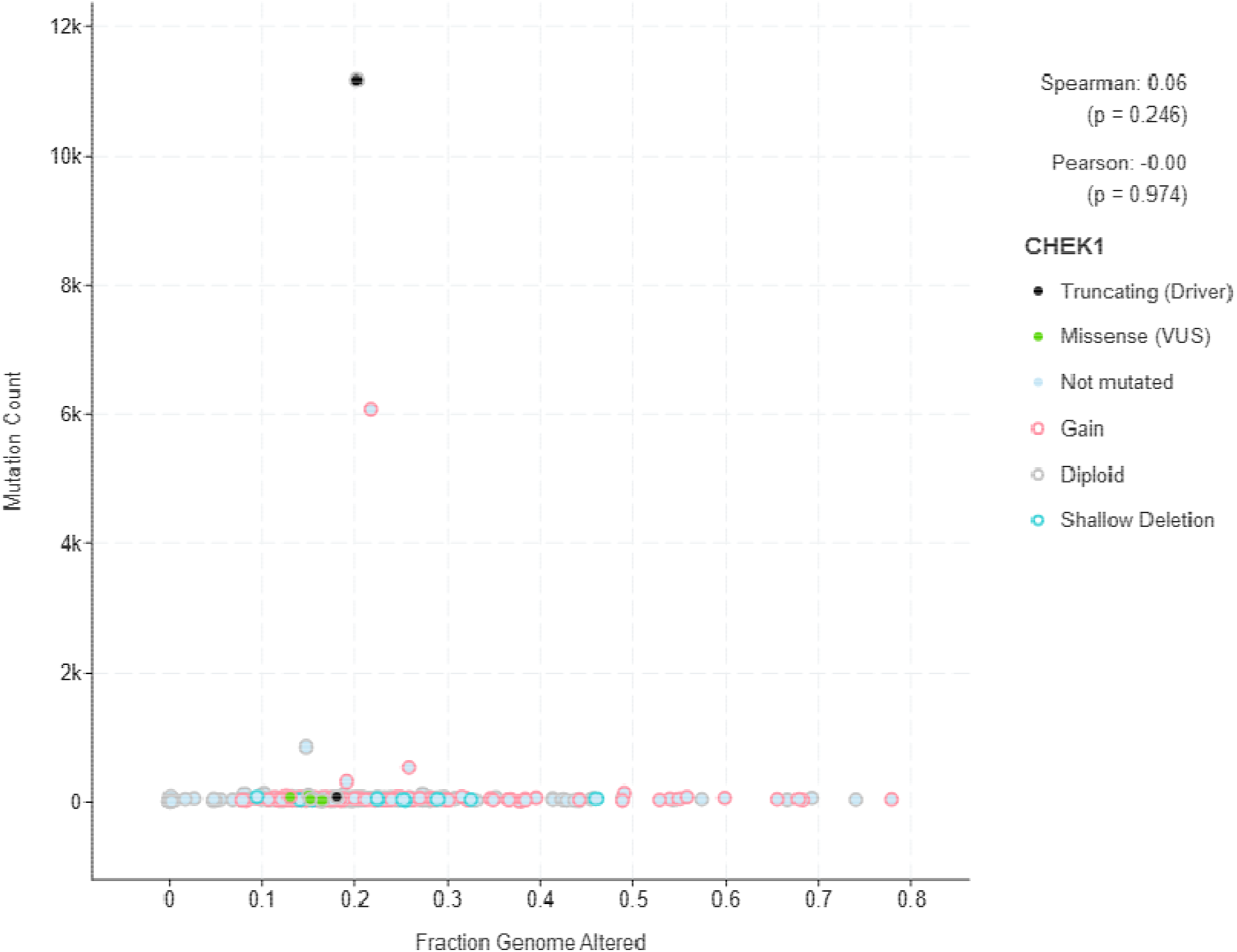
Scatter plot showing mutation count versus fraction of genome altered, stratified by CHEK1 status in TCGA glioblastoma (GBM). Scatter plot shows the association between total mutation count and fraction of genome altered in TCGA glioblastoma (GBM) samples, stratified by *CHEK1* mutation and copy number status. Each point represents an individual tumor sample, colored by *CHEK1* alteration type (truncating mutation, missense variant of uncertain significance, gain, diploid, or shallow deletion). Unlike lower-grade gliomas, *CHEK1* status in GBM shows no significant correlation with either mutation burden or genomic instability (Spearman ρ = 0.06, *p* = 0.246; Pearson *r* ≈ 0, *p* = 0.974). These findings suggest that in advanced disease, replication stress and chromosomal chaos are pervasive and decoupled from *CHEK1* alterations, reflecting widespread genomic instability and checkpoint pathway saturation characteristic of GBM and late-stage gliomas.

Table 1 shows co-occurrence analysis of key genomic alterations in TCGA lower-grade glioma (LGG). This table summarizes the frequency with which major molecular alterations occur together across the TCGA LGG cohort, along with odds ratios and statistical significance. ATRX and TP53 mutations display a highly significant co-occurrence (odds ratio > 3, *q* < 0.001), reflecting their well-established cooperation in the astrocytoma molecular subtype. CHEK1 alterations also show a statistically significant, though more modest, co-occurrence with TP53 (*odds ratio 1*.*248, q* = 0.043), suggesting that replication stress signaling is preferentially activated in TP53-deficient tumors. Together, these findings underscore the strong molecular linkage between ATRX and TP53 in glioma pathogenesis and highlight CHEK1 as a potential therapeutic target in this genetic context. q is the Benjamin Hochberg false discovery rate [10].

**Table 1.** Co-occurrence analysis of key genomic alterations in TCGA lower-grade glioma (LGG). This table summarizes the frequency with which major molecular alterations occur together across the TCGA LGG cohort, along with odds ratios and statistical significance. ATRX and TP53 mutations display a highly significant co-occurrence (odds ratio > 3, *q* < 0.001), reflecting their well-established cooperation in the astrocytoma molecular subtype. CHEK1 alterations also show a statistically significant, though more modest, co-occurrence with TP53 (*odds ratio 1,248, q* = 0.043), suggesting that replication stress signaling is preferentially activated in TP53-deficient tumors. Together, these findings underscore the strong molecular linkage between ATRX and TP53 in glioma pathogenesis and highlight CHEK1 as a potential therapeutic target in this genetic context. Benjamin Hochberg false discovery rate (q-value).

| A | B | Neither | A Not B | B Not A | Both | Log2 Odds Ratio | p-Value | q-Value | Tendency |
| --- | --- | --- | --- | --- | --- | --- | --- | --- | --- |
| ATRX | TP53 | 1278 | 43 | 444 | 248 | >3 | <0.001 | <0.001 | Co-occurrence |
| CHEK1 | TP53 | 1366 | 13 | 708 | 16 | 1.248 | 0.028 | 0.043 | Co-occurrence |

Table 2 shows co-occurrence of *CHEK1* and *IDH2* alterations in TCGA glioblastoma (GBM). This table summarizes the frequency with which *CHEK1* and *IDH2* alterations occur independently or together in GBM, along with calculated odds ratios, *p*-values, and *q*-values (Benjamini–Hochberg false discovery rate). Although *IDH2* alterations are infrequent in GBM, they significantly co-occur with *CHEK1* amplification, suggesting that replication stress signaling and metabolic reprogramming are functionally linked even in highly aggressive tumors. The enrichment of *CHEK1*/*IDH2* co-alterations identifies a rare but biologically distinct GBM subset that may remain dependent on CHK1-mediated DNA damage response pathways and could therefore benefit from replication stress–targeted therapeutic strategies.

**Table 2.**
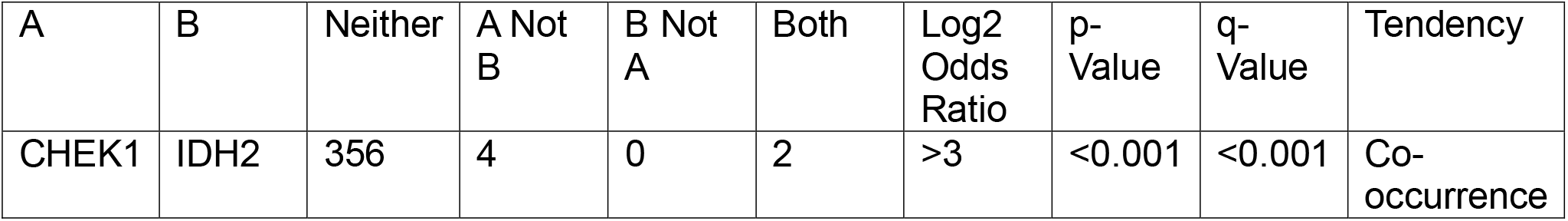
Co-occurrence of *CHEK1* and *IDH2* alterations in TCGA glioblastoma (GBM). This table summarizes the frequency with which *CHEK1* and *IDH2* alterations occur independently or together in GBM, along with calculated odds ratios, *p*-values, and *q*-values (Benjamini–Hochberg false discovery rate). Although *IDH2* alterations are infrequent in GBM, they significantly co-occur with *CHEK1* amplification, suggesting that replication stress signaling and metabolic reprogramming are functionally linked even in highly aggressive tumors. The enrichment of *CHEK1*/*IDH2* co-alterations identifies a rare but biologically distinct GBM subset that may remain dependent on CHK1-mediated DNA damage response pathways and could therefore benefit from replication stress–targeted therapeutic strategies.

| A | B | Neither | A Not B | B Not A | Both | Log2 Odds Ratio | p-Value | q-Value | Tendency |
| --- | --- | --- | --- | --- | --- | --- | --- | --- | --- |
| CHEK1 | IDH2 | 356 | 4 | 0 | 2 | >3 | <0.001 | <0.001 | Co-occurrence |

## Discussion

Our study identifies *CHEK1* amplification and overexpression as central features of ecDNA-driven glioma biology, linking replication stress signaling to genomic instability, tumor aggressiveness, and clinical outcomes. By systematically analyzing TCGA datasets, we demonstrate that high *CHEK1* activity is strongly associated with poor survival—particularly in *1p/19q*-non-codeleted lower-grade gliomas—and correlates with hallmarks of chromosomal chaos, including elevated mutation burden and a higher fraction of genome altered. These findings underscore *CHEK1*’s dual role as a biomarker of replication stress and a therapeutic vulnerability in gliomas shaped by ecDNA dynamics.

An intriguing observation in our analysis is that unlike in lower-grade glioma where *CHEK1* amplification strongly predicts poor outcome, in GBM lower *CHEK1* copy number is paradoxically associated with worse survival (Figure 2). This apparent contradiction likely reflects fundamental differences in tumor biology between early- and late-stage disease. In GBM, replication stress and CHK1 pathway activation are nearly universal due to widespread ecDNA amplification and high oncogenic load, resulting in a checkpoint saturation state in which additional *CHEK1* copies confer little selective advantage. Moreover, aggressive GBM subclones may evolve alternative mechanisms to tolerate replication stress or bypass CHK1 dependence altogether, such as alterations in ATR signaling, fork protection pathways, or enhanced tolerance of DNA damage. These adaptations can enable unchecked proliferation despite low *CHEK1* expression and are often associated with treatment resistance and highly aggressive clinical behavior. Additionally, total ecDNA burden in GBM may decouple from *CHEK1* status, with tumors harboring amplifications of other oncogenes (*EGFR, PDGFRA, MDM2*) exhibiting severe genomic instability independent of *CHEK1*. Thus, reduced *CHEK1* copy number in GBM may serve as a surrogate marker of advanced clonal evolution and checkpoint independence, rather than a sign of diminished aggressiveness.

An alternative explanation is that ecDNA leads to rapid, non-gradual evolution and drug resistance. The GBM finding, therefore, might not be a contradiction but an artifact of the pervasive genomic instability inherent in late-stage disease. The finding of no significant correlation between CHEK1 status and instability in GBM (Figure 5) suggests CHEK1’s role as a *marker* is saturated and has been superseded by other, perhaps compensatory, mechanisms like ATR signaling. This complex, late-stage biology highlights the need for functional studies, as correlation alone fails to capture the true therapeutic window.

The co-occurrence of *CHEK1* and *IDH2* alterations in GBM (Table 2), though relatively uncommon, provides additional insight into the molecular complexity of advanced gliomas and reveals a potential convergence point between metabolic reprogramming and replication stress signaling. *IDH2* mutations are classically associated with epigenetic remodeling, altered NADPH metabolism, and the production of oncometabolites such as 2-hydroxyglutarate, which can profoundly influence DNA repair and chromatin state [12]. Their co-alteration with *CHEK1* suggests that tumors harboring these changes may exploit both metabolic and replication stress–adaptation mechanisms to sustain proliferation under adverse conditions. Importantly, this dual dependency could create a distinct therapeutic vulnerability: while most GBMs evolve beyond reliance on single pathways, *CHEK1*–*IDH2* co-altered tumors may remain particularly sensitive to replication stress–targeted therapies or metabolic–checkpoint inhibitor combinations. These findings highlight the value of integrating *IDH2* status into biomarker-driven stratification strategies and suggest that a subset of GBM patients may derive greater clinical benefit from approaches that simultaneously target metabolic and DNA damage response pathways.

The unique biology of ecDNA provides a mechanistic explanation for the observed dependence on CHK1. Unlike chromosomal amplifications, ecDNA replicates and segregates stochastically, driving rapid, non-Mendelian evolution and profound intratumoral heterogeneity. These circular DNA elements frequently carry oncogenes (*EGFR, MYC*), immune-modulatory genes, and regulatory elements that boost transcriptional output. However, their intense transcriptional activity, occurring simultaneously with DNA replication, creates replication–transcription conflicts. These collisions generate replication stress, characterized by stalled forks, double-strand breaks, and activation of the DNA damage response (DDR). CHK1, as a key mediator of the ATR-CHK1 pathway, is indispensable in resolving such conflicts and maintaining replication fork stability [1].

Our results suggest that *CHEK1* amplification is not merely a passenger event but reflects selective pressure to sustain proliferation in the context of ecDNA-induced stress. Tumors with *CHEK1* amplification show significantly higher genomic instability, suggesting that elevated CHK1 activity may be required to tolerate the heightened DNA damage landscape of ecDNA-rich cancers. This model is consistent with recent preclinical work showing that CHK1 inhibition disproportionately affects ecDNA-positive tumor cells by exacerbating replication stress beyond a tolerable threshold, leading to catastrophic DNA damage and cell death.

Our analysis also reveals that the prognostic significance of *CHEK1* in LGG is strongly influenced by *1p/19q* co-deletion status. Tumors harboring *1p/19q* co-deletion— hallmarks of oligodendroglioma [13]—display significantly lower *CHEK1* expression and improved survival, even when copy number gains are present. This suggests that these tumors experience less replication stress and genomic instability, possibly reflecting fundamental differences in ecDNA generation or propagation. By contrast, *1p/19q*-intact astrocytomas show higher *CHEK1* expression and worse outcomes, consistent with their greater reliance on CHK1-mediated checkpoint signaling.

These findings support a model in which the molecular context of glioma modulates the degree to which replication stress contributes to tumor behavior and therapeutic vulnerability. Stratifying patients by *1p/19q* status could therefore enhance the predictive value of *CHEK1* as a biomarker and improve the precision of replication stress–targeted therapies.

Our findings have several important therapeutic implications. First, they support the rationale for CHK1 inhibition as a targeted approach in gliomas with high *CHEK1* activity or genomic instability. Preclinical models have demonstrated that blocking CHK1 selectively kills ecDNA-positive tumor cells while sparing normal cells, suggesting a potential therapeutic window. Early-phase clinical trials with CHK1 inhibitors have shown proof-of-concept efficacy, although toxicity and pharmacokinetic limitations remain significant challenges [1].

Second, combination strategies may be necessary to maximize efficacy and minimize adverse effects. For example, co-inhibition of CHK1 and ribonucleotide reductase (RNR)—which supplies the dNTP building blocks for DNA synthesis—has shown synergistic activity in preclinical models, overwhelming the replication machinery and preventing ecDNA repair and propagation [14]. Other approaches under investigation include targeting mitotic machinery essential for ecDNA segregation, exploiting synthetic lethal interactions in the DDR network, and modulating transcriptional stress responses.

Finally, *CHEK1* may serve as a biomarker for patient selection in clinical trials. Measuring *CHEK1* expression or copy number, along with genomic instability metrics such as mutation burden or FGA, could help identify tumors most likely to respond to replication stress–targeted therapies. Integration of *CHEK1* status into molecular diagnostics could therefore refine therapeutic decision-making and guide personalized treatment strategies.

Despite these advances, several key questions remain. Most critically, our analysis is correlative and does not directly demonstrate that ecDNA-driven replication stress causes *CHEK1* dependency. Functional studies—including perturbation of CHK1 in ecDNA-high versus ecDNA-low tumor models—will be essential to validate causality. Additionally, methods for direct ecDNA quantification (e.g., AmpliconArchitect, Circle-Seq, or long-read sequencing) should be integrated into future studies to better link ecDNA burden with replication stress phenotypes.

The therapeutic landscape remains complex. Although CHK1 inhibition is promising, early clinical trials have encountered dose-limiting toxicities and pharmacological barriers. Future efforts must focus on optimizing drug design, improving blood–brain barrier penetration, and identifying synergistic drug combinations. Moreover, the intricate network of DDR pathways may necessitate multiplexed targeting strategies rather than monotherapy.

The clinical implications of our findings are substantial. As a central mediator of replication stress tolerance, *CHEK1* represents both a prognostic biomarker and a potential therapeutic target in gliomas characterized by ecDNA-driven genomic instability. Assessment of *CHEK1* expression and copy number—alongside molecular subtype markers such as *IDH* mutation and *1p/19q* co-deletion—could be readily incorporated into existing diagnostic workflows using standard genomic platforms, enabling risk stratification and personalized treatment planning. Patients with high *CHEK1* activity, particularly those with *1p/19q*-non-codeleted astrocytomas, may benefit from clinical trials testing CHK1 inhibitors or rational drug combinations that exacerbate replication stress. Moreover, integrating *CHEK1* status with measures of mutation burden and chromosomal instability could serve as a predictive biomarker to identify responders to emerging replication stress–targeted therapies. Ultimately, translating these insights into clinical practice could improve patient selection, enhance therapeutic efficacy, and pave the way for precision oncology strategies that exploit the unique vulnerabilities of ecDNA-driven gliomas.

### Weaknesses and Limitations

Limitations include the observational nature of our analysis, lack of direct ecDNA quantification, and the absence of functional validation.

The most critical limitation is the lack of direct ecDNA quantification. The current study infers ecDNA activity and dependency through proxies (CHEK1 expression/amplification and genomic instability). While this correlation is significant, the conclusion that CHEK1 is a target in ecDNA-driven gliomas remains an inference. High CHEK1 could be driven by other forms of replication stress unrelated to ecDNA.

As a purely observational analysis of public data (TCGA), the study cannot establish causation. It identifies associations, but it cannot functionally validate whether inhibiting CHK1 *causes* tumor death in specific subtypes, or whether CHEK1 simply *marks* an aggressive phenotype.

CHK1 inhibition is in early trials, but there is a crucial, cautionary context: early clinical trials for CHK1 inhibitors have already faced setbacks due to toxicity and poor pharmacokinetics. The scientific promise is clear, but the clinical path is difficult [1].

## Conclusion

In summary, our findings position *CHEK1* as a central node in the interplay between ecDNA biology, replication stress, and genomic instability in gliomas. CHEK1 amplification and overexpression are tightly linked to aggressive tumor behavior, particularly in *1p/19q*-non-codeleted astrocytomas, and may define a biologically distinct subset of gliomas with heightened vulnerability to replication stress–targeted therapies. By integrating *CHEK1* status into molecular stratification frameworks and advancing combination treatment strategies, it may be possible to exploit this vulnerability and overcome one of the most formidable barriers to effective glioma therapy—its relentless genomic adaptability.

## Data Availability

All data produced are available online at
https://xenabrowser.net
https://www.cbioportal.org

https://www.cbioportal.org

